# A Comprehensive HPV-STI NGS Assay for Detection of 29 HPV Types and 14 non-HPV Sexually Transmitted Infections

**DOI:** 10.1101/2021.11.24.21266783

**Authors:** Zhihai Ma, Baback Gharizadeh, Xingsheng Cai, Mengzhen Li, María Dolores Fellner, Jorge Alejandro Basiletti, Rita Mariel Correa, María Celeste Colucci, Gabriela Baldoni, Martín Vacchino, Patricia Galarza, María Alejandra Picconi, Chunlin Wang

## Abstract

Sexually transmitted infections (STIs) are prevalent throughout the world and impose a significant burden on individual health and public health systems. Missed diagnosis and late treatment of STIs can lead to serious complications such as infertility and cervical cancer. Although sexually transmitted co-infections are common, most commercial assays target one or a few STIs. The HPV-STI ChapterDx Next Generation Sequencing (NGS) assay detects and quantifies 29 HPVs and 14 other STIs in a single-tube and single-step PCR reaction and can be applied to tens to thousands of samples in a single sequencing run. The assay was evaluated in this study, and the limit of detection was 100% at 50 copies for all targets, and 100%, 96%, 88% at 20 copies for 34, 8, and 1 target, respectively. The performance of this assay has been compared to Roche cobas HPV test, showing an overall agreement of 97.5% for hr-HPV, and 98.5% for both, HPV16 and HPV18. The assay also detected all HPV-infected CIN2/3 with 100% agreement with Roche cobas HPV results. Moreover, several co-infections with non-HPV STIs, such as *C. trachomatis, T. vaginalis, M. genitalium*, and HSV2 were identified. The ChapterDx HPV-STI NGS assay is a user-friendly, easy to automate and cost-efficient assay, which provides accurate and comprehensive results for a wide spectrum of HPVs and STIs.

## INTRODUCTION

Sexually transmitted infections (STIs) are prevalent globally and World Health Organization (WHO) estimates that annually there are 376 million new cases of four curable STIs: *Chlamydia trachomatis, Neisseria gonorrhoeae, Treponema pallidum* (syphilis), and *Trichomonas vaginalis*. Prevalence of several viral STIs is similarly high with an estimated 500 million people infected with herpes simplex type 2 (HSV2), and approximately 291 million women harboring human papillomaviruses (HPV) (1). STI prevalence varies by region, gender. STI epidemics cause a profound impact on physical and psychological health worldwide [1, 2].

HPVs are a large group of viruses with more than 230 completely characterized types, and new HPV types being continuously found [3]. Among them, 40 types known to infect human anogenital tract are grouped into high risk (hr)-HPV and low risk (lr)-HPV types based on their oncogenic potential [4, 5]. Hr-HPVs are the causative agents of cervical cancer, and have been detected in 99.7% of cervical cancers [6, 7]. Fourteen hr-HPV types are considered to cause most cervical cancers worldwide. Nevertheless, other hr-types have also been reported to cause cervical cancer [4, 5]. HPV oncogenic potential is highly type-dependent as some types are more oncogenic. Many studies have shown that extended genotyping information holds important clinical value for patient management and triage of HPV positive women [8-10].

HPV viral load quantification has been shown to be a predictor of infection persistence or clearance [11]. It has also been reported that HPV viral load is associated with risk of persistent infection and precancer [12-15], indicating viral load as a candidate risk marker [16]. Currently, viral load is measured mainly by quantitative polymerase chain reaction (qPCR) on a limited number of hr-HPV types such as HPV16 and HPV18. An assay that can detect and quantify many HPVs in a single-tube PCR reaction is a needed tool for association studies of viral load and precancer.

Besides HPVs, several bacteria and viruses infect human anogenital area. Earlier studies indicated that non-HPV STIs could facilitate the entry of HPV virions into host cells, change in the immunological response pathways, and decrease the host’s ability to clear HPV infection [16-18]. Co-infections between hr-HPVs and non-HPV STIs such as *Chlamydia trachomatis, Neisseria gonorrhoeae, Trichomonas vaginalis, Ureaplasma urealyticum*, and HSV2 have been reported to be associated with HPV persistence, cervical dysplastic and neoplastic lesion [19-26]. Co-infections between HPVs and non-HPV STIs are widespread. Since most commercial kits detect one or two STIs, multiple tests are required for comprehensive STI identification.

This work presents the evaluation of the ChapterDx HPV-STI NGS assay (NGS assay in short) that amplifies 29 HPVs and 14 STIs (Table 1) with species/type-specific primers in a single-tube and single-step PCR reaction followed by NGS (Fig. 1). This assay can detect and quantify these 43 targets from tens to thousands of samples in a single Illumina sequencing run (depending on the sequencing platform and sequencing kit).

**Table 1.** The list of 43^*^ HPV/STI in the NGS panel

|  |  |  |
| --- | --- | --- |
| 19 HR-HPV Types | HPV-16, 18, 26, 31, 33, 35, 39, 45, 51, 52, 53, 56, 58, 59, 66, 68a, 68b, 73, 82 |  |
| 10 LR-HPV Types | HPV-6, 11, 40, 42, 43, 44, 55, 61, 81, 83 |  |
| 14 STI Species/Types | *Chlamydia trachomatis* (serovars A to K) | *Ureaplasma urealyticum* |
|  | *Neisseria gonorrhoeae* | *Ureaplasma parvum* |
|  | *Mycoplasma genitalium* | Herpes simplex virus type 1 |
|  | *Trichomonas vaginalis* | Herpes simplex virus type 2 |
|  | *Treponema pallidum* | Varicella zoster virus |
|  | *Haemophilus ducreyi* | *Chlamydia trachomatis* serovar L1 |
|  | *Mycoplasma hominis* | *Chlamydia trachomatis* serovar L2 |

\*For simplicity, HPV68a and HPV68b are listed as separate HPV types; *Chlamydia trachomatis* serovar L1 and L2 are listed as separate STIs.

**Fig 1.**
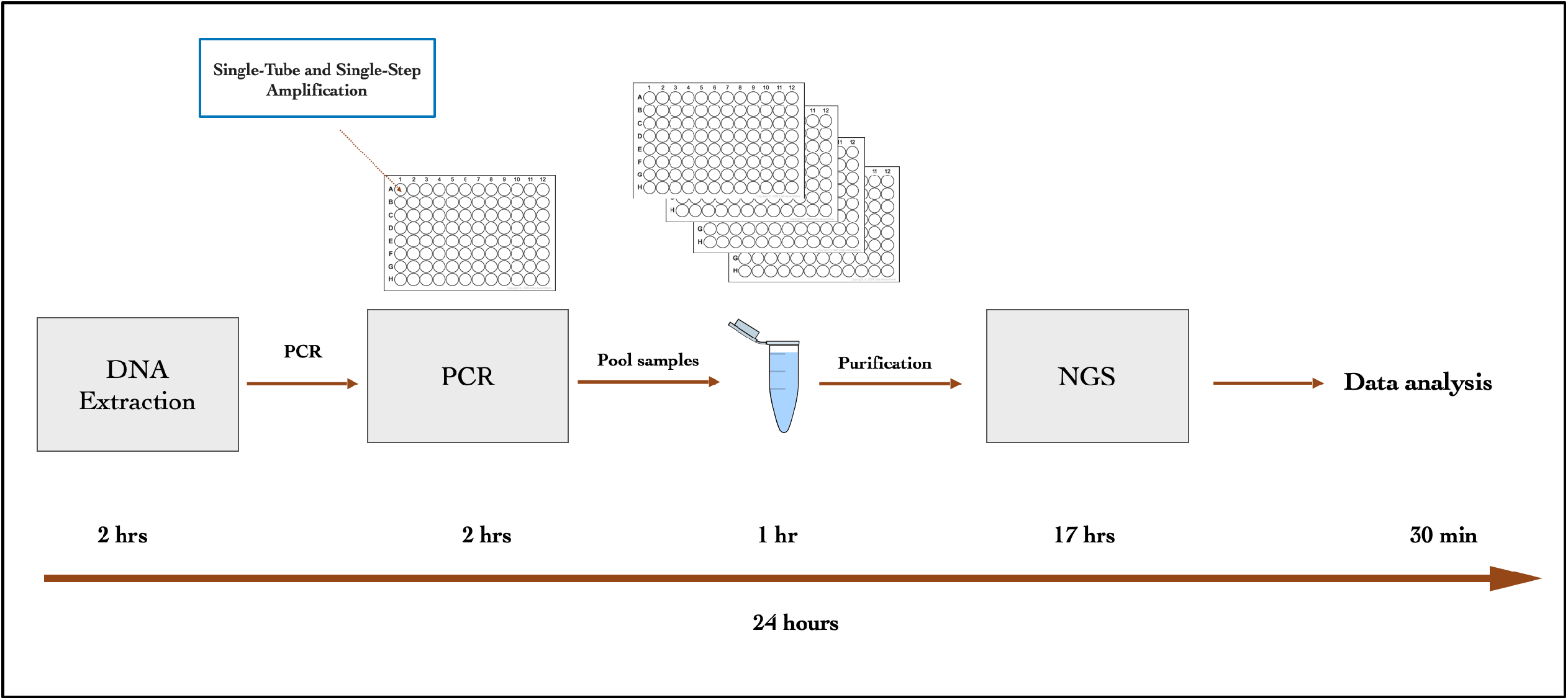
A schematic illustration of ChapterDx HPV-STI NGS workflow

## MATERIALS AND METHODS

### Clinical HPV screening specimens

A cohort of 274 blind samples that had previously been analyzed by conventional cytology/histology and Roche cobas HPV Test (Roche, Pleasanton, CA, USA) was selected for this study. Within this cohort, 102, 101 and 57 samples were negative for intraepithelial lesions or malignancy (NILM), low-grade squamous intraepithelial lesion (LSIL) and highgrade squamous intraepithelial lesion (HSIL), respectively; and five were cancer and nine samples had no cytology results. The available histological results for this cohort were: cervical intraepithelial neoplasia grade 1 (CIN1) (N = 4), CIN2 (N = 1) and CIN3 (N = 58), 3 squamous cell carcinoma (SCC) and 2 adenocarcinoma (AdC). HPV samples were shipped from Oncogenic Viruses Service, National and Regional HPV Reference Laboratory, National Institute of Infectious Diseases-ANLIS “Dr. Malbrán”, Buenos Aires, Argentina to Chapter Diagnostics in Menlo Park, California, USA, for comprehensive HPV-STI analysis.

### ChapterDx HPV-STI NGS kit

ChapterDx HPV-STI NGS kit targets 19 hr-HPVs, 10 lr-HPVs, 14 non-HPV STIs, and GAPDH as internal control. Table 1 shows the HPV genotypes and non-HPV STIs covered by the kit.

### Controls for analytical performance

A set of 43 synthetic control DNA fragments for all targets were synthesized by Integrated DNA Technologies (Coralville, IA). They were pooled and diluted into 100, 50 and 20 copies per vial in a final volume of 5 µl. Each pool was spiked with 10 ng human genomic DNA. The pools were amplified and sequenced in 23 replicates for 100 copies and 24 replicates for 50 and 20 copies to assess the limit of detection (LoD) of the NGS assay. The acceptable consensus for an assay’s LoD at the lowest concentration is ≥95% for at which 19/20 replicates are positive [27].

### DNA extraction and PCR

Genomic DNA extraction for 274-sample cohort were performed by Roche cobas HPV Test (cobas for short) platform. For genotyping 75-sample cohort, DNA extraction was performed by Qiagen EZ1 virus kit (Qiagen, Hilden, Germany). All the specimens were measured by Qubit 3 (ThermoFisher, CA, USA) to monitor the presence and concentration of DNA prior to PCR. One-step multiplex PCR was performed in 15 µl final volume in a 96-well plate on a Veriti thermocycler (ThermoFisher, CA, USA). The PCR reaction consisted of target-specific primers (29 HPV, 14 STI and internal control), barcoded universal primers, sample DNA, DNA polymerase, dNTPs and PCR buffer. The PCR conditions were according the ChapterDx HPV-STI NGS kit instructions.

### Library preparation and next-generation sequencing

After PCR, the amplified products were pooled into a 1.5 ml tube (or a 15 ml tube depending on the sample size). A portion of the pooled amplicons was purified with SPRIbeads (Beckman Coulter, CA, USA) according to the manufacturer’s instructions. The purified pooled amplicons concentration was measured on a Qubit fluorometer, and the concentration was adjusted for sequencing according to Illumina library preparation instructions. The library was sequenced on an Illumina MiniSeq platform using an Illumina Mid-Output sequencing kit (Illumina, CA, USA). The workflow is depicted in Figure 1.

### Sequence data analysis software and interpretation

1) Decoding: sequencing reads (FASTQ format) for forward and reverse reads and forward and reverse indexing reads are input in ChapterDx Analysis Software. The software assigns sequencing reads for a sample based on the sequence of both forward and reverse index reads with no mismatch; 2) Mapping: sequencing reads are mapped onto reference sequences using the Smith-Waterman algorithm with options as nucleotide match reward is 1, nucleic mismatch penalty is -3, cost to open a gap is 5, and cost to extend a gap is 2. Only the alignment of best match is kept for each sequencing read if the alignment score is beyond 60. Alignments of paired reads with identical reference sequences are kept. Reads are labeled to the reference based on their corresponding alignments; 3) Genotyping: a sample is called positive for a species/type when there are more reads than the preset cutoff reads labelled with the corresponding reference.

### NGS detection threshold

Based on ChapterDx HPV-STI NGS sequence data from WHO LabNet international proficiency HPV panel [28], the threshold for positive samples were set at 3 reads. Samples below the threshold were considered negative.

### Statistical analysis

The HPV and STI species/types frequencies, cytology and histology were calculated by Microsoft excel. Overall agreement for 14 hr-HPV, HPV16, 18 and other 12 HPVs between ChapterDx HPV-STI NGS and Roche cobas HPV assays were calculated by Cohen’s Kappa coefficients with 95% confidence intervals (CIs). The genotyping comparison between ChapterDx HPV-STI NGS and Genomica Clart assays were calculated by Microsoft excel.

## RESULTS

### Analytical performance of the assay

To evaluate the analytical performance and sensitivity of the assay, a set of 43 synthetic control DNA fragment pools of 100 copies (23 replicates), 50 copies (24 replicates), 20 copies (24 replicates) and 0 copies (blank) spiked with 10 ng human genomic DNA were analyzed to determine the LoD. 100 copies and 50 copies control DNA were detected in all replicates. For the 20 copies pool replicates, the detection rate was 100% (24/24) for 34 targets, 95.8% (23/24) for HPV6, 11, 39, 42, 43, 45, 56 and 83 (8 targets) and 87.5% (21/24) for *Mycoplasma genitalium* (Table 2a and 2b). The lowest concentration LoD is recommended at ≥95% (for 19/20 replicates) [27], therefore, the LoD was determined at 20 copies for all targets except for *Mycoplasma genitalium*. There was no sequencing read for the blank sample.

**Table 2a.** LoD results of 43 control fragment pools for 23 replicates of 100 copies and 24 replicates of 50 and 20 copies

| Copy Number (43 control Pool) | Type/Species | Replicate Detection Rate | % LoD Detection |
| --- | --- | --- | --- |
| 100 Copies | 43/43 | 24/24 | 100% |
| 50 Copies | 43/43 | 24/24 | 100% |
| 20 Copies | 34/43 | 24/24 | 100% |
|  | 8/43 | 23/24 | 95.8% |
|  | 1/43 | 21/24 | 87.5% |

**Table 2b.** 20 copies LoD for 9 control fragments below100% agreement

| Species/Types | Replicate agreement | LoD agreement |
| --- | --- | --- |
| HPV6 | 23/24 | 96% |
| HPV11 | 23/24 | 96% |
| HPV39 | 23/24 | 96% |
| HPV42 | 23/24 | 96% |
| HPV43 | 23/24 | 96% |
| HPV45 | 23/24 | 96% |
| HPV56 | 23/24 | 96% |
| HPV83 | 23/24 | 96% |
| *M. Genitalium* | 21/24 | 88% |

### Agreement between ChapterDx HPV-STI NGS assay and Roche cobas HPV Test in cervical samples with cyto-histological diagnosis

To evaluate the agreement with the Roche cobas HPV test, 274 clinical samples were analyzed by ChapterDx HPV-STI NGS assay. Out of 274 samples, the results for 267 samples agreed with those by cobas HPV test for 14 hr-HPVs. Among those 267 samples, 241 were positive and 26 were negative for 14 hr-HPVs. The overall agreement for 14 hr-HPVs between both methods was 97.5% with kappa 0.867. The CIN1 (n=4), CIN2/3 (n=59) were positive for hr-HPV by both cobas and the NGS assays (Table 3). Of the 5 cancer samples, one squamous carcinoma sample and one adenocarcinoma sample were negative for 14 hr-HPV by both cobas and the NGS assay. The adenocarcinoma sample was positive for HPV44, 81 by the NGS assay, which was confirmed by reverse line blot assay [29].

**Table 3.**
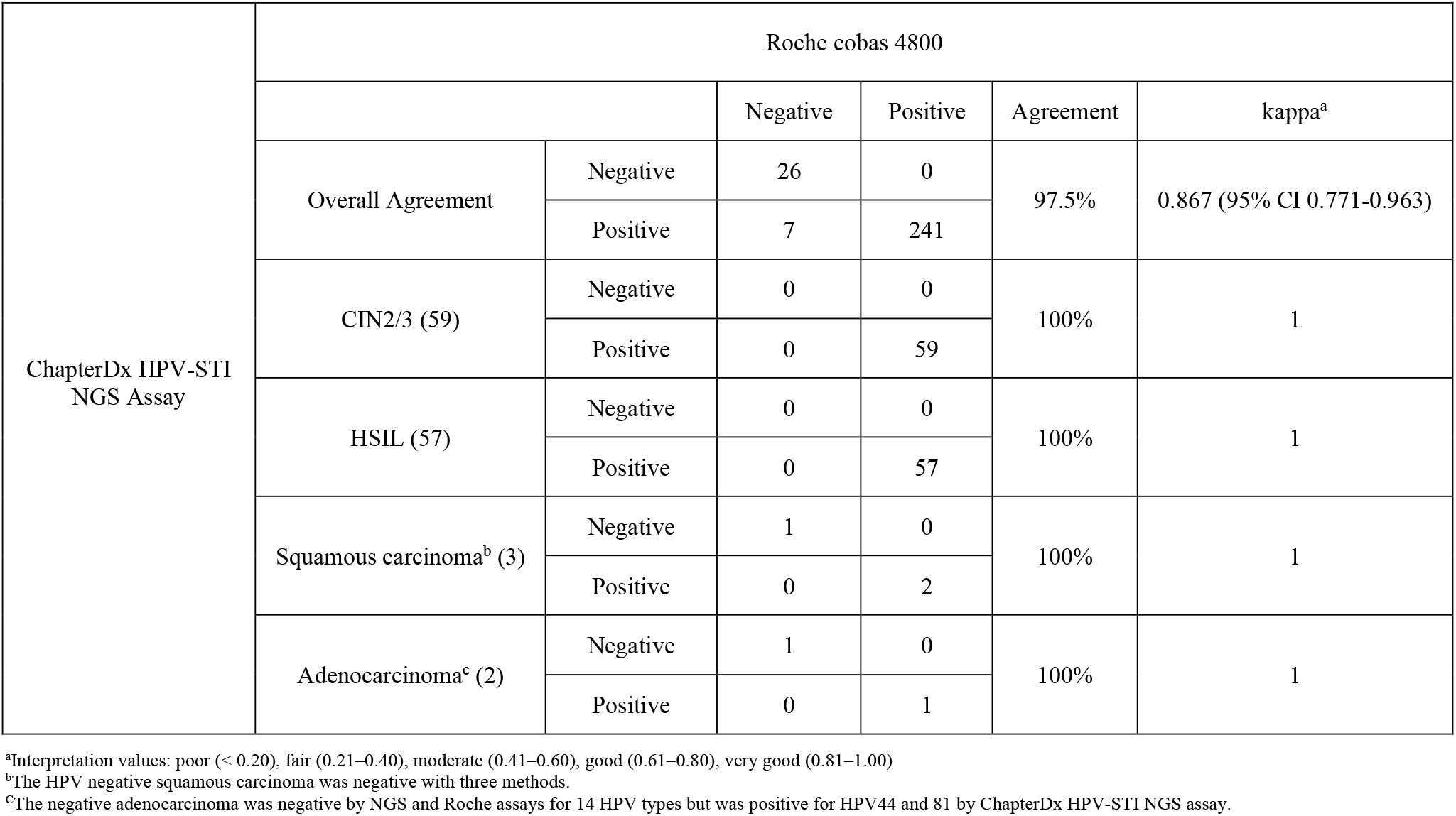
Overall concordance of 274 clinical samples between ChapterDx HPV-STI NGS assay and Roche cobas for 14 high-risk HPVs, stratified by histology and cytology

The overall agreement for HPV16 between cobas and the NGS assays was 98.5% (270/274) with a kappa value of 0.967 (Table 4a). Two of the HPV16 samples were scored negative. However, one and two HPV16 reads were detected in each of these two samples, respectively. The overall agreement for HPV18 was 98.5% (70/274) with a kappa value of (0.901) and the overall agreement for other HPVs was 96% (263/274) with kappa value of 0.906. For the other HPVs, two HPV samples were also detected by the NGS assay as HPV52 and HPV31/52, respectively, but were below the NGS detection threshold and scored negative. Of the 29 HPV targets in the NGS panel, 28 were detected among the samples analyzed (except for HPV73). In this cohort, co-infections between 2 and 12 HPVs were detected in 146 samples (shown in Table 4b).

**Table 4a.**
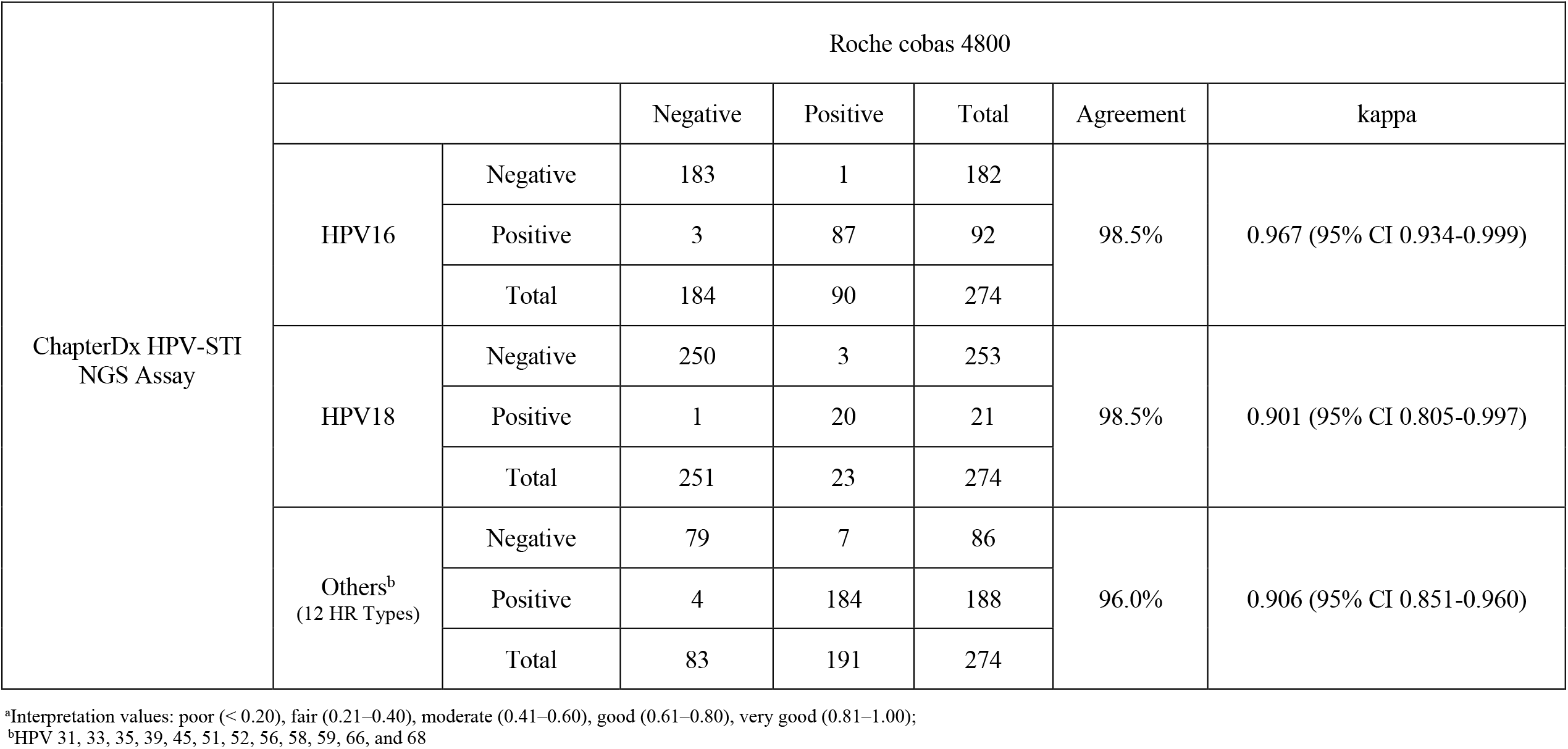
Concordance between ChapterDx HPV-STI NGS assay and Roche cobas assay for 14 high-risk HPVs

**Table 4b.** Nr of 2 to 12 HPV co-infections in 274 samples

| Nr of HPV co-infections | 2 | 3 | 4 | 5 | 6 | 7 | 8 | 9 | 10 | 12 | total |
| --- | --- | --- | --- | --- | --- | --- | --- | --- | --- | --- | --- |
| Nr of specimens | 67 | 29 | 20 | 19 | 5 | 1 | 1 | 2 | 1 | 1 | 146 |

In the 274-sample cohort, ChapterDx HPV-STI NGS assay detected a total frequency of 205 non-HPV STIs, of which *U. parvum* (32.1%) and *M. hominis* (25.2%) comprised the majority of the non-HPV STIs (Table 5a). 52.1% (143/274) of the samples were positive for 7 out of 14 non-HPV STIs, harboring a range of 1-4 STIs per sample (Table 5b). HPV and non-HPV STIs co-infections were observed in 135/274 (49.3%) of the samples. Table 6 lists results by Roche cobas assay and the NGS assay for 9 samples with co-infections of multiple HPVs and non-HPV STIs, demonstrating detection of HPV and non-HPV STIs by the NGS assay.

**Table 5a.** Other STIs detected in the 274 samples by ChapterDx HPV-STI NGS assay

| STIs detected | Nr/% |
| --- | --- |
| *Chlamydia trachomatis* | 4 (1.5%) |
| *Trichomonas vaginalis* | 16 (5.8%) |
| *Mycoplasma genitalium* | 1 (0.36%) |
| *Mycoplasma hominis* | 69 (25.2%) |
| *Ureaplasma parvum* | 88 (32.1%) |
| *Ureaplasma urealyticum* | 23 (8.4%) |
| Herpes simplex virus type 2 | 4 (1.5%) |

**Table 5b.** Nr of 2 to 4 non-HPV STI co-infections in 274 samples

| Nr of STI co-infections | 2 | 3 | 4 | Total |
| --- | --- | --- | --- | --- |
| Nr of specimens | 45 | 6 | 1 | 52 |

**Table 6.** Comparison of ChapterDX HPV-STI NGS and Roche cobas for samples with combined HPV and STI co-infection

| Sample | Roche Cobas | Chapter Diagnostics HPV-STI NGS Panel |
| --- | --- | --- |
| 1 | HPV16 and others | HPV16, 35, *Trichomonas vaginalis*, *Mycoplasma hominis* |
| 2 | Others | HPV33, 56, 43, *Trichomonas vaginalis*, *Mycoplasma hominis* |
| 3 | HPV16, 18, others | HPV16, 18, 31, 33, 45, 52, 56, 68a, 42, 61 *Trichomonas vaginalis*, *Mycoplasma hominis* |
| 4 | Others | HPV58 and *Chlamydia Trachomatis* |
| 5 | Others | HPV16, 39, and Herpes Simplex II (HSV2) |
| 6 | Others | HPV66, 68a and *Chlamydia Trachomatis* |
| 7 | Others | HPV31, 51, *Mycoplasma genitalium*, *Trichomonas vaginalis*, *Mycoplasma hominis*, *Ureaplasma Parvum* |
| 8 | HPV16 and others | HPV16, 45, 11, 81, 83, Herpes Simplex II (HSV2), *Mycoplasma hominis*, *Trichomonas vaginalis* |
| 9 | Others | HPV58, 42, *Chlamydia trachomatis* |

In this study, 192 positive samples were sequenced per Illumina Mid-Output sequencing kit on Illumina MiniSeq platform. Table S1 lists the estimated number of samples for different Illumina sequencing platforms and kits.

## Discussion

In the present study, the performance of ChapterDx HPV-STI NGS assay was compared to Roche cobas assay. Forty-three control DNA pools of 100 and 50 copies were detected in all replicates and for 20 copies replicates, *M. genitalium* had an LoD below 95%. The lower detection rate for *M. genitalium* could be due to synthetic error in control DNA, which affects amplification efficiency. In this study, HPV52 control DNA fragment had to be re-synthesized due to multiple errors found in primer regions.

The NGS and cobas assays showed a high overall agreement, which indicates a good performance of the NGS assay on a set of well-studied samples; nevertheless, a larger study is required based on Meijer’s criteria [30] for further clinical evaluation. In this 274-sample cohort, none of the discrepant results involved HSIL, CIN1, CIN2/3 or cancer samples. The adenocarcinoma sample that was negative for 14 hr-HPVs by both methods, harbored HPV44, 81, which are lr-HPV types and not considered carcinogenic. Due to type-specific amplification, the NGS assay was able to detect a range of 2-12 HPV coinfections in 146/274 of samples, allowing unbiased analysis of existing HPV types in each sample.

Earlier studies have shown evidence of association between hr-HPVs and non-HPV STIs (including commensal STIs) with cervical carcinogenesis [17-26, 31]. In this study, 52.1% samples in the 274-sample cohort harbored non-HPV STIs, of which 87.8% were commensal infections of three species: *M. hominis, U. urealyticum* and *U. parvum*. Moreover, almost half of the samples carried co-infections of HPV and non-HPV STIs. Thus, a comprehensive assay detecting multiple HPV and non-HPV STIs in a single test, is a useful tool to diagnose and study the correlation of hr-HPV and non-HPV co-infections. It is worth mentioning that the 14 non-HPV STIs in the NGS assay have different pathogenic characteristics and some of them are considered commensal species of the genital tract (such as *M. hominis, U. urealyticum* and U. *parvum*), existing in both healthy and symptomatic individuals [32].

HPV genotyping has been mostly focused on HPV16 and HPV18 due to their high prevalence in cervical cancer, although, other hr-types indicate high positive predictive value (PPV) [8, 10]. Earlier studies suggested that extended HPV genotyping provides a better risk stratification and identification of women at increased risk of cervical cancer by simply providing individual risk assessment of HPV positive women [33-35]. The results of the two cohorts in this study show that co-infections of multiple HPVs and non-HPV STIs are widespread (Table 5). Due to limitation of current widely used assays, many of those STIs can go undetected or more than one assay might be needed.

It has been reported that non-transient hr-HPV infections and high viral loads is a risk marker for dysplasia and carcinoma in situ in normal cytology [11-16]. Such studies have been typically focused on few high-risk types. The NGS assay uses type-specific HPV primers for amplification and generates quantification information for each type in the panel. The wide spectrum of STIs and their quantification information could be useful for studying the association between STIs and diseases.

There are several NGS-based HPV assays [36-38] that commonly use more than one round of PCR. ChapterDx HPV-STI NGS assay uses one-step amplification workflow, which significantly reduces time/labor, the risk of cross contamination and allows easier automation.

## Conclusion

ChapterDx HPV-STI NGS Assay is cost-effective as it detects a much broader spectrum of HPV/STI than other commercial kits, at a similar or lower cost per sample, with proven analytical performance; therefore, it is a valuable tool for large epidemiology and surveillance studies. Once evaluated with larger, well-characterized cohorts for clinical validation, this assay could also be applied to cervical cancer screening programs.

## Data Availability

All data produced in the present study are available upon reasonable request to the authors.

## Ethics approval and consent to participate

This cohort is part of a screening study that is being carried out by the INEI-ANLIS Malbrán with the collaboration of the “Prof. A. Posadas” Hospital (Argentina); it was approved by the Research Ethics Board and the Ethics Committee of this hospital. Written informed consent was obtained before enrolling the participant women, and confidentiality and anonymity were guaranteed.

## Authors’ contributions

Z.M., B.G., and C.W. are involved in the overall study of this assay and manuscript writing. X.C. and M. Li participated in the LoD assay and manuscript writing. The INEI-ANLIS Malbrán investigators are responsible for the selection of clinical samples previously characterized, participate in the analysis, interpretation of the data and revision of the manuscript.

---

**Supplementary Table S1.**
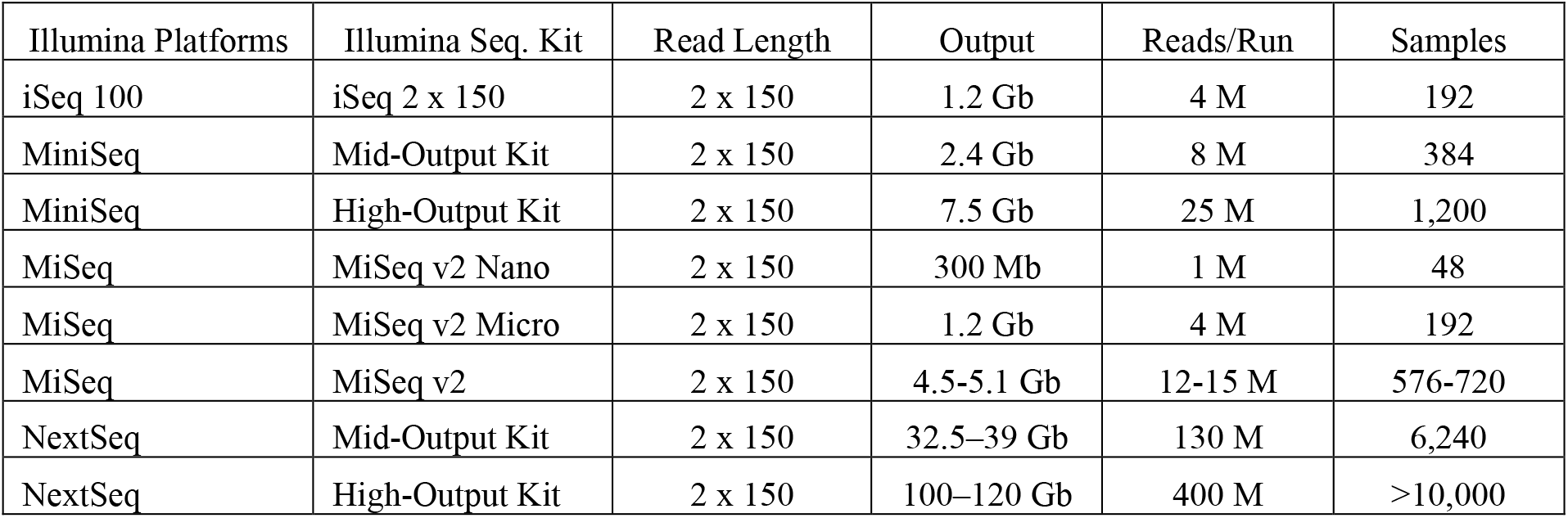
Comparison of Illumina sequencing platforms and their respective sequencing kits for HPV-STI analysis

| Illumina Platforms | Illumina Seq. Kit | Read Length | Output | Reads/Run | Samples |
| --- | --- | --- | --- | --- | --- |
| iSeq 100 | iSeq 2 x 150 | 2 x 150 | 1.2 Gb | 4 M | 192 |
| MiniSeq | Mid-Output Kit | 2 x 150 | 2.4 Gb | 8 M | 384 |
| MiniSeq | High-Output Kit | 2 x 150 | 7.5 Gb | 25 M | 1,200 |
| MiSeq | MiSeq v2 Nano | 2 x 150 | 300 Mb | 1 M | 48 |
| MiSeq | MiSeq v2 Micro | 2 x 150 | 1.2 Gb | 4 M | 192 |
| MiSeq | MiSeq v2 | 2 x 150 | 4.5-5.1 Gb | 12-15 M | 576-720 |
| NextSeq | Mid-Output Kit | 2 x 150 | 32.5–39 Gb | 130 M | 6,240 |
| NextSeq | High-Output Kit | 2 x 150 | 100–120 Gb | 400 M | >10,000 |

